# COMPARATIVE STUDY ON THE PREVALENCE OF MALNUTRITION AMONG PUBLIC PRIMARY SCHOOL PUPILS IN RURAL AND URBAN COMMUNITIES IN WARRI SOUTH

**DOI:** 10.1101/2022.12.23.22283888

**Authors:** Chinwendu Sandra Emeagi, Uchechukwu Ifeanyichukwu Apugo

## Abstract

**Background:** Malnutrition among primary school-age children has continued to pose a major public health issue, especially in developing countries. Nigeria is ranked amongst the top ten countries with the highest prevalence of undernutrition in children while about 2,300 children die daily in Nigeria as a result of malnutrition.

**Aim:** To determine and compare the prevalence of malnutrition among public school pupils in rural and urban communities in Warri South LGA.

**Materials and Method:** This school-based comparative cross-sectional study made use of a bio-data form that collected socio-demographic details and anthropometric measurements of the pupils. The nutritional status of the pupils was obtained using WHO AnthroPlus software, while the resulting data were analysed and presented accordingly.

**Results:** A total of 330 pupils (165 each from urban and rural public primary schools) were recruited, out of which more of the pupils in urban areas were younger, had parents who were better educated and employed while those from rural areas had higher family sizes. However, there was no difference in the sex and type of family of the pupils. The prevalence of malnutrition was reported as 36.7% with the prevalent form of malnutrition discovered as underweight (35.0%), followed by stunting (13.9%), overweight/obese (8.5%), and wasting (7.2%). Wasting, stunting, and overweight were higher among urban pupils, while the rate of underweight was higher among rural children.

**Conclusion:** This high prevalence of undernutrition among urban pupils could be attributed to poor nutrition arising from a myriad of interrelated circumstances such as poor feeding, eating practices, and recurrent infection. Hence, the need for improved sensitisation of mothers concerning correct childhood feeding and provision of balanced nutrition.

## INTRODUCTION

Childhood malnutrition to date remains a major global public health problem that has constituted a long-time barrier to a healthy life and a constant threat to human capital development.^1^ This silent emergency has been defined by the World Health Organization (WHO) as the cellular imbalance between the supply of nutrients and energy and the body’s demand for them to ensure growth, maintenance, and specific functions,^2^ or the consumption of dietary nutrients either insufficiently or excessively.^3^ Hence, it refers not only to deficiency states but also to excess or imbalance in the consumption of calories, proteins, and/or other nutrients.^4^ Therefore, it represents a spectrum that can exist as micronutrient malnutrition, under-nutrition, and over-nutrition or obesity,^5-7^ though it has been used over time to attribute to or often used interchangeably with under-nutrition in explaining nutritional problems.^8^

Infants and young children are the most vulnerable to malnutrition due to their high nutritional requirements for physical growth and development,^9^ as their age represents a dynamic period when children undergo mental, emotional, and social change.^10^ According to Babatunde et al.^11^, malnutrition affects all aspects of children’s lives in a gruesome manner by resulting in poor academic performance, absenteeism, reduced intellectual achievement, delayed cognitive development morbidity, and mortality. Hence, children who are malnourished often fail to thrive, as well as more likely to suffer from impaired physical and intellectual growth which makes them less productive during adulthood.^12^

Globally, malnutrition among primary school-age children, especially those in developing countries, is a major public health concern,^13^ especially among children in sub-Saharan Africa (SSA) and Southern Asian countries.^6,14^ The Joint Child Malnutrition estimates of UNICEF, WHO, and World Bank in 2017, approximated that about 155 million children (22.9%) are stunted and about 52 million children (7.7%) are wasted.^15^ Also, the mortality rate due to malnutrition in developing countries is about 41% in the age group of 6 – 60 months, while more than one-third of under-five deaths in LMICs are attributable to undernutrition.^16^ According to Ocheke and Thandi,^17^ Nigeria is ranked among the top ten countries with the highest prevalence of underweight (too thin for age), stunting (too short for age), and wasting (too thin for height) in children. Also, UNICEF reported that about 2,300 children die daily, while one in seven children in developing countries will die before attaining school age because of malnutrition.^18^ The recent National Demographic and health survey (NDHS) report also showed that 50% of childhood mortality is caused by malnutrition, while nearly 2 in 5 (37%) children under five in Nigeria are still stunted, 7% of them are wasted and 22% were underweight.^19^

Several studies have identified various factors that affect children’s nutritional status. These factors include poverty, lack of exclusive breastfeeding, unsatisfactory or inadequate food intake, poor consumption of vitamin supplements or fortified foods, large family size, poor sanitation, large family size, food insecurity and safety, social inequality, poor sanitation, poor maternal nutrition during pregnancy, severe and repeated infections/illnesses.^9,12,20^ The NDHS report also showed that children whose mothers have no education (54%) and those from the poorest households (55%) were most likely to be stunted while rural children were identified to have higher levels of stunting, wasting, and underweight, compared to urban children.^19^ Similarly, Khan, Khan & Raza^21^ and Abdulahi et al.^22^ reported that rural-urban differentials in the place of residence of children also influence nutritional outcomes among children. This is because children raised in urban areas are generally healthier (in terms of nutritional outcomes) than their rural counterparts.^5,23^ Another area of consideration is that of lifestyle and dietary factors as consumption of energy-dense and fatty diet from fast food outlets, reduced active commuting to school, and use of energy-saving devices (computer games and television programs) have been associated with malnutrition in the area of over nutrition.^24^ Hence, this study is set at comparing the prevalence of malnutrition among public primary school pupils in Warri South Local Government Area (WSLGA) of Delta state, South-south, Nigeria.

## METHODOLOGY

### Study Design and setting

This school-based comparative cross-sectional study was conducted in Warri South Local Government Area of Delta state, Nigeria. The LGA represents one of the three LGAs that make up the Warri Federal constituency (Warri South-West and Warri North), all of which have a preponderance of government-owned (public) primary schools. During the 2019/2020 academic year, Asoro^25^ identified that there are 1,113 public primary schools in Delta state. However, according to the record of the statistics division of the state Ministry of Education, there are 54 public primary schools in WSLGA, with a population of 20,655 pupils (the third highest in the state) and a teacher strength of 1235. Hence there is a 1:17 teacher-to-pupil ratio.

### Study Population

The target population for this study consisted of apparently healthy pupils, aged 6 – 12 years, registered in public primary schools within the rural and urban areas of WSLGA.

### Sample Size Determination and Sampling Techniques

A sample size of 236 (118 for rural and urban each) was determined using the double proportion comparison formula^26^; n = (Z_α/2_+Z_β_)^2^(p_1_(q_1_) +p_2_(q_2_)) / (p_1_-p_2_)(p_1_-p_2_), and considering: 95% confidence interval, a power of 80%, 5% margin of error, 27% and 45% prevalence of stunting among Children in urban and rural areas respectively,^19^ and a 10% non-response rate.

The selection of the participants was based on multi-staged sampling which first identify the public primary schools in the federal constituency using the list of schools in the constituency, stratified them into urban and rural primary schools, randomly selected a total of 4 primary schools from each area, followed by random selection of the arms per each grade by balloting and lastly, a simple random selection of pupils from the selected arms

### Study Instruments

This study made use of a bio-data form which was divided into two parts. Part A collected information from all eligible pupils such as age (date of birth verified from school register), sex, parents’ educational and employment status, type of family, and size of family, while part B was used to record the measurement of the weight and height of the pupils.

The measurements of the weight and height of the pupils were obtained from each pupil according to the protocol described by WHO.^28^ Their weights were taken with the aid of an electronic weighing scale (Camry Model Number BR9011) in accordance with Ivanovic et al.^29^ and this was done in the morning (immediately after assembly), as it is known that there are diurnal variations in weight.^24^ The height or stretch stature was determined in accordance with Ivanovic et al.^30^ and Eze et al.^31^ with the aid of a portable height measure (Seca® stadiometer). The weight and height were converted to nutritional indices: weight-for-age *z*-score (WAZ), height-for-age *z*-score (HAZ), and body mass index-for-age Z-scores (BAZ) which were obtained using the WHO AnthroPlus software for measuring malnutrition in school-age children and adolescent aged 5 – 19 years,^7,32,32^ and the result used to obtain the nutritional status of the pupils

### Statistical Analysis

The anthropometric data obtained from this study was calculated by application of the WHO growth references and analysed with IBM Statistical Package for Social Sciences Software (SPSS vs 23). The results were expressed as percentages and presented in tables and charts. Descriptive statistics were used to determine the frequency and standard deviations (SDs) of the anthropometric measurements while differences in means across the two areas (urban and rural) were compared using Student’s t-test.

### Ethical Consideration

Ethical approval was obtained from the Ethical Review Committee of the University of Port Harcourt. Official permission was obtained from the Delta state ministry of education before carrying out this study. The Head teachers at the schools were duly informed of the nature of the study and their permission was granted while written informed consent was obtained permission from the parents of the pupils. The pupils also gave assent and were informed of their right to withdraw from the study at any point.

## RESULTS

### Socio-demographic characteristics of Primary school pupils and their Parents

The result presented in Table 1 below shows that there is a statistically significant (p < 0.05) difference between the age of the pupils in the urban and rural areas as more of the children in the urban schools were younger with 20.6% aged 7 years, in comparison with the rural pupils who appeared to be older with 26.7% aged 10 years. On the other hand, the result of the analysis of the sex of the pupils revealed no statistically significant (p > 0.05) difference between the urban and rural pupils as 50.9% of the entire pupils (urban = 50.3% and rural = 51.5%) were males and 49.1% (urban = 49.7% and rural = 48.5%) females. The result also showed that there was a statistically significant (p < 0.05) difference with respect to the educational level and occupational status of the parents of the pupil, as the parents of the urban pupils, were more educated (tertiary = 53.3%) and employed (23.6%) in comparison with the parents of the rural pupils who had mostly primary school education (38.8%) and involved in trading (82.4%). Also, the result of the type of family of the pupil did not show any statistically significant (p > 0.05) difference between the urban and rural pupils, though there was a statistically significant (p < 0.05) higher family among the pupils in the rural areas.

**Table 1:** Age and sex pupils and characterizes of their parents

| Variables (n=330) | Urban (n=165)<br>n (%) | Rural (n=165)<br>n (%) |
| --- | --- | --- |
| **Age category** |  |  |
| 6 | 7 (4.2) | 8 (4.8) |
| 7 | 34 (20.6) | 17 (10.3) |
| 8 | 25 (15.2) | 26 (15.8) |
| 9 | 33 (20.0) | 7 (4.2) |
| 10 | 22 (13.3) | 44 (26.7) |
| 11 | 26 (15.8) | 22 (13.3) |
| 12 | 18 (10.9) | 41 (24.8) |
|  | *Chi Square = 39.286; p-value = <0.0001\** |  |
| **Sex** |  |  |
| Male | 83 (50.3) | 85 (51.5) |
| Female | 82 (49.7) | 80 (48.5) |
|  | *Chi Square = 0.924; p-value = 0.826* |  |
| **Educational level of parent** |  |  |
| None | 9 (5.5) | 47 (28.5) |
| Primary | 24 (14.5) | 64 (38.8) |
| Secondary | 44 (26.7) | 47 (28.5) |
| Tertiary | 88 (53.3) | 7 (4.2) |
|  | *Chi Square = 113.130; p-value = <0.0001\** |  |
| **Occupational status of parent** |  |  |
| Farmer | 10 (6.1) | 24 (14.6) |
| Artisan (traders) | 105 (63.6) | 136 (82.4) |
| Employed | 39 (23.6) | 3 (1.8) |
| Unemployed | 11 (6.7) | 2 (1.2) |
|  | *Chi Square = 46.840; p-value = <0.0001\** |  |
| **Type of family** |  |  |
| Nuclear | 155 (93.9) | 158 (95.8) |
| Extended | 10 (6.1) | 7 (4.2) |
|  | *Chi Square = 0.558; p-value = 0.455* |  |
| **Size of family** |  |  |
| Three | 17 (10.3) | 13 (7.9) |
| Four | 63 (38.2) | 39 (23.6) |
| Five or more | 85 (51.5) | 113 (68.5) |
|  | *Chi Square = 10.140; p-value = 0.006\** |  |

\* Statistically significant ( $p < 0.05$ ).

### Nutritional status of pupils in Urban and Rural Public Schools

Table 2 above presents the result of the prevalence of the different malnutrition indices among pupils in urban and rural areas. According to the table, more of the pupils were underweight (35.0%), while wasting (7.2%) was the least reported form of malnutrition among the pupils. Comparison of the result among the pupils in the urban and rural areas revealed that wasting and stunting were more prevalent among urban school children (15.4% and 20.0% respectively) thereby showing a statistically significant (p < 0.05) difference in comparison with the pupils in the rural areas. On the other hand, the prevalence of underweight was higher among rural children (51.7%) in comparison to those in the urban areas (26.3%) while overweight/obese was seen to be prevalent among those in the urban area (11.1%). However, this did not show any statistically significant (p > 0.05) difference.

**Table 2:**
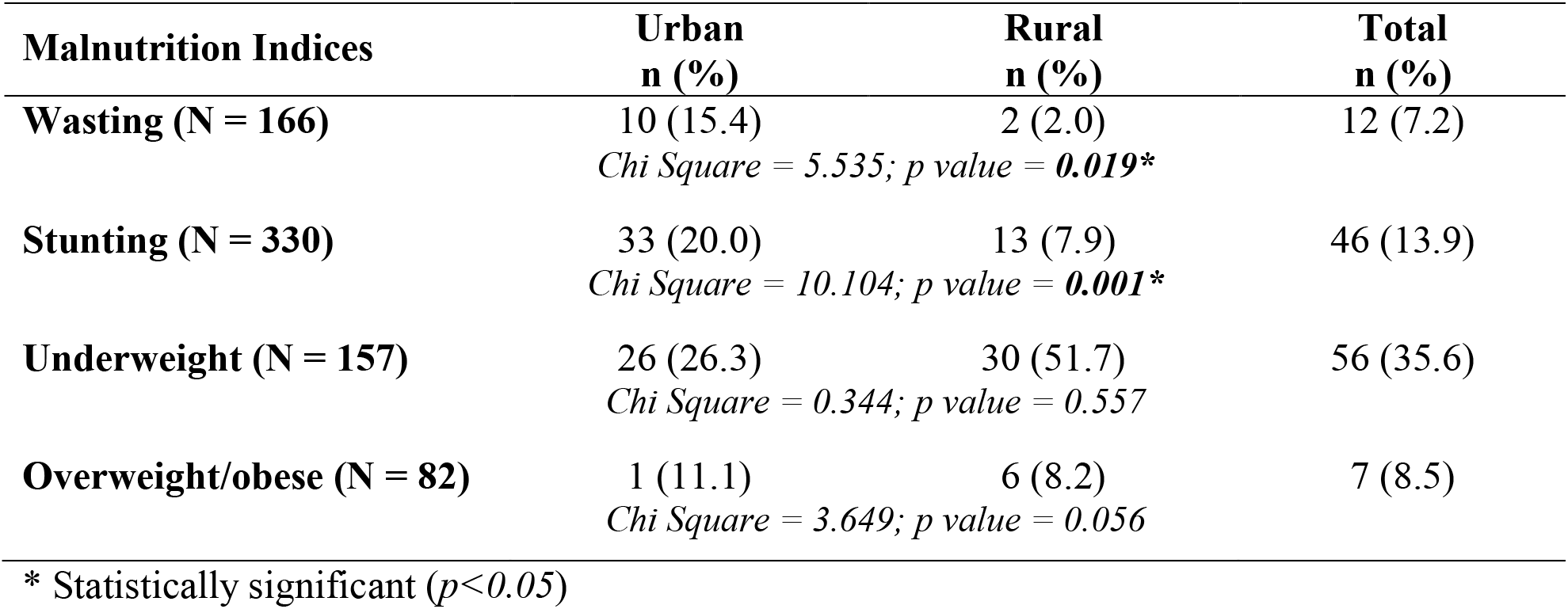
Prevalence of malnutrition among public primary school pupils in WSLGA

### Prevalence of malnutrition between pupils in Urban and Rural Public Schools

The number of malnourished and healthy pupils is presented in figure 1 above. According to the result, comparison across the urban and rural divide showed only slight differences as 42.4% and 30.9% of the urban and rural pupils respectively were seen to be malnourished while 57.6% and 69.1% of the urban and rural pupils respectively were seen to be healthy. This shows that a total of 209 (63.3%) pupils are healthy while 121 (36.7%) were reported to be malnourished. Though the urban pupils were slightly more malnourished and slightly less healthy than the rural pupils, there was no statistically significant (p > 0.05) difference in the malnutrition status between the urban and rural public primary school pupils.

**Figure 1:**
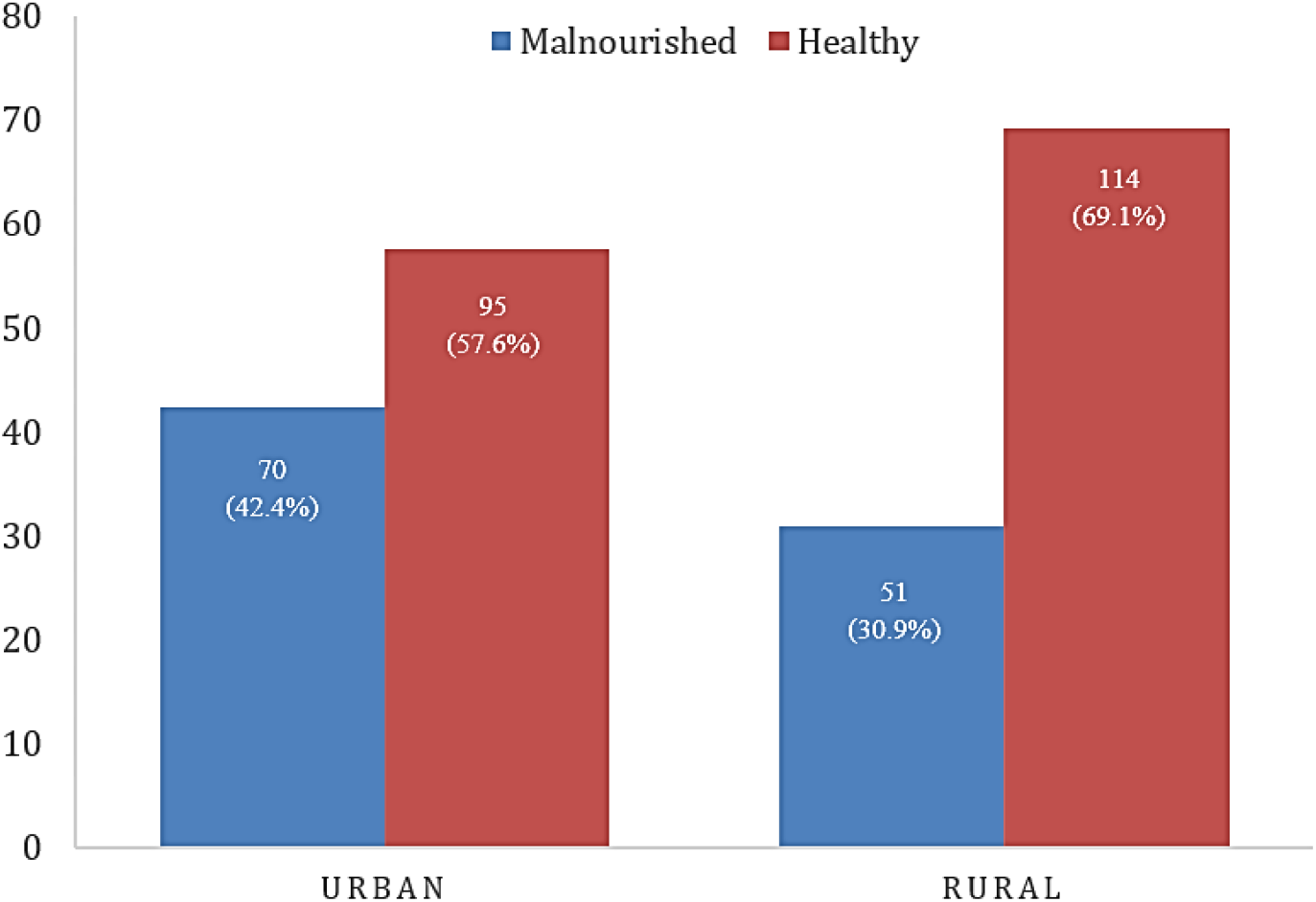
malnourished and healthy pupils in public primary schools in WSLGA

## DISCUSSION

A total of 330 pupils (165 each from the urban and rural public primary schools) were recruited for this study. The determination of their age showed that pupils in urban areas were younger, and had parents who were better educated and employed while those from rural areas had higher family sizes. There was no difference in the sex and type of family of the pupils. The older age of the pupil in the rural schools could be linked to late schooling either due to poverty as a result of low educational attainment, income, or occupational status of parents while the younger age of the urban pupil could be as a result of the occupational status, availability of finance and high educational attainment of the parents. Children of employed parents tend to start schooling early so their parents can return to work. Furtherance to this, the parents also understand the importance of education.

The result of the nutritional status of the pupils revealed that underweight (35.0%) was the most prevalent form of malnutrition, followed by stunting (13.9%), overweight/obese (8.5%), and wasting (7.2%). Though there is currently no national or state prevalence data for malnutrition among the school-aged population for comparison, the 2018 NDHS report for children under five years reported a lower level of underweight (22%) and obese/overweight (2%), but a higher rate of stunting (37%) and a similar rate for wasting (7%).^19^ In relation to other studies, the rate of undernutrition in this study is higher than that of Akor et al.^34^ which reported lower rates of underweight (10.3%), stunting (11.1%), and wasting (2.4%), while Saltzman et al.^35^ reported a higher prevalence of stunting (15%) but lower underweight (13%). In more recent studies, Hassan et al.^27^ and Terefe et al.^36^ reported a higher prevalence of stunting (34.2% and 31.8% respectively), but a lower prevalence of wasting (0.9% and 2.5%), and underweight (3.4% and 4.7%) respectively, among school-aged children. Also, the prevalence of wasting (7.2%) which often indicates recent and severe weight loss due to lack of adequate quality and quantity of food and/or presence of frequent or prolonged illnesses, is lower than the 14% and 19.4% in Ethiopia,^37^ and Ghana,^38^ respectively, as well as the 33.3% and 50.4% in India,^39^ and Sri Lanka^40^ respectively. The prevalence of stunting, which is generally due to inadequate feeding and/or the presence of infectious diseases which lasted relatively over a long period or recurred many times,^41^ as recorded in this study, is lower than that of Roba, Abdo & Wakayo^42^ (37%) but higher than that of Anurag et al.^39^ (1.7%). Again, in this study, the prevalence of underweight which is largely a measure of previous and recent weight loss or failure to gain weight,^43^ is seen to be higher in comparison with the findings of Roba, Abdo & Wakayo^39^ (14.9%) and Mwaniki and Makokha^40^ (14.66 %), but similar with that of Sarma, Wijesinghe and Sivananthawerl,^40^ in Sri Lanka (33.7%). These differences might be due to the average family expenditure for food, socio-economic status, food habit frequency, hygienic condition, genetics, and the difference in culture and geographical conditions.^44^ Also, the majority of the children used in the study are said to be of lower socioeconomic classes despite the fact that half were residing in an urban area, while most of the children may be more engaged in more physically demanding activities, such as long treks to school. Also, Adewale et al.^35^ added that a high rate of undernutrition among urban pupils could arise due to the consequence of poor nutrition arising from multifaceted and interrelated circumstances such as poor feeding, eating practices, and repeated infection, while the low rate of overweight/obesity as reported in this study may be explained by the fact that overnutrition is only recently emerging as a public health problem in our society unlike in the developed world where it has taken full effect due to rapid modernization, changes in feeding and more sedentary lifestyles.^46^

The comparison of the nutritional status of the two groups showed that the prevalence of wasting, stunting, and overweight was higher among the urban pupils (15.4%, 20.0%, and 11.1% respectively) in comparison with those who school in the rural areas (2.0%, 7.9%, and 8.2% respectively), while the rate of underweight was higher among the rural children (51.7%) in comparison with the urban (26.3%). Similar to this report, two studies conducted in urban and rural India, documented a higher prevalence of wasting and stunting in the urban areas (33.3% and 18.5% respectively) than in the rural areas (12.3% and 9.2% respectively), ^47,48^ while Aziz and Devi^49^ reported that incidence of stunting is more among children from urban areas in Malaysia. Also, the studies of Veghari^50^, Bello, Ekekezie and Afolabi^51^ and Umeokonkwo et al.^7^ still in agreement with this report showed that the prevalence of underweight was higher among pupils residing in the rural areas in comparison with their urban colleagues. On the contrary, several studies have reported that undernutrition (underweight, stunting, and wasting) is the major health problem among rural public primary school children in comparison with urban children.^46,52,53^ Some other disagreeing reports include that of Davis et al.^54^ which showed that the prevalence of obesity was higher among rural children, while Maddah et al.^55^ observed that most of the children in urban areas in Zahedan, Iran were underweight. Onabanjo et al.^56^ also reported that more of the children population from the rural LGAs in their study were stunted, compared to their urban counterparts.

The relationship between the malnourished (29.7%) and healthy (70.3%) pupils showed no significant difference across the two areas. However, in comparison with previous studies, it was revealed that the finding of Abebe et al.^57^ in Ethiopia and Sarma, Wijesinghe & Sivananthawerl^40^ in Sri Lanka reported a higher rate of malnutrition (34.6% and 60.2% respectively). Though the prevalence of malnutrition is not high in this study, its existence is suggestive of long-term nutritional deprivation in the children,^7^ while the disparity could be due to a difference in the composition of diets with essential nutrients like proteins, carbohydrates, fats, vitamins, and minerals. Also, contrary to widely held notions that malnutrition is due to poverty, Torpy^27^ suggested that it may be caused by people choosing to eat the wrong types of food, rather than a lack of what to eat, while Adewale et al.^45^ added that it could be as a result of repeated exposure to poor sanitation and interactive effects of poor energy, nutrient intake, and infection.

## Conclusion and Recommendation

This present study has shown that the prevalence of malnutrition (36.7%) in the LGA is higher among urban pupils, with underweight (35.0%) reported as the prevalent form of malnutrition in the LGA. Comparing the finding across the two groups showed that wasting, stunting, and overweight was higher among the urban pupils, while the rate of underweight was higher among the rural children. Based on the findings of high prevalence of malnutrition (mostly undernutrition) observed in this study, it is necessary to engage parents in the study area in sensitization programs geared towards educating them on child nutrition and how to provide their wards with balanced nutrition, as a means of promoting nutritional health for the pupils.

## Data Availability

All data produced in the present study are available upon reasonable request to the authors

